# Extracting smoking history from clinical notes for lung cancer screening decision support: comparing a structured-judgment model with general-purpose large language models

**DOI:** 10.64898/2026.09.24.26363906

**Authors:** Adam Wright, Siru Liu, Aileen Wright

**Affiliations:** Department of Biomedical Informatics, Vanderbilt University Medical Center, Nashville, TN, USA

**Keywords:** clinical decision support, lung cancer screening, natural language processing, large language models, smoking, benchmark

## Abstract

**Objective:** To compare the accuracy, cost, and speed of a low-cost, non-generative structured-judgment model and four general-purpose large language models (LLMs) for extracting smoking status, pack-years, and quit date from clinical notes to support lung cancer screening (LCS) clinical decision support (CDS).

**Materials and Methods:** We built a synthetic, shareable benchmark of 3,000 outpatient notes in three conditions: 1,000 template-generated notes, 1,000 realistic, “messy” notes written by an LLM from structured facts, and 1,000 “messy” notes that required complex arithmetic to determine pack-years and quit dates. Ground truth reference labels were programmatically generated before each note was created. We compared TypeSafe Jev 1.13 with Claude Haiku 4.5, Claude Sonnet 5, GPT-6 Luna and GPT-6 Sol, each using an identical structured output schema. We determined United States Preventive Services Task Force (USPSTF) 2021 and American Cancer Society (ACS) 2023 eligibility from each system’s output in code.

**Results:** Jev made the correct eligibility decision for 99.1%, 99.9%, and 94.4% of notes in the three conditions, compared with 97.8%, 98.4%, and 98.1% for Haiku, 100.0%, 99.6%, and 99.8% for Sonnet, 99.7%, 98.8%, and 98.5% for Luna, and 100.0%, 99.8%, and 99.6% for Sol. In the complex condition, Jev produced 27 false positive and 14 false negative screening flags per 1,000 notes, the most of any system. Luna cost $0.12 to $0.21 per 1,000 notes and Jev $0.61 to $0.64, compared with $2.44 to $4.22 for Sol, $3.37 to $4.43 for Haiku, and $3.76 to $6.61 for Sonnet. Jev was the fastest system (median 0.45 to 1.21 seconds per note, compared with 1.71 to 3.50 seconds for the others).

**Discussion:** All models tested had high accuracy. The structured-judgment model had the lowest latency, which could allow for synchronous use in clinical decision support systems rather than batched, cached invocation. The cost of the structured-judgment model was low, but the newest cost-efficient LLM, Luna, was less expensive and more accurate on the complex notes, though its latency was higher. Accuracy, latency, and cost are all moving quickly as new models are released.

**Conclusion:** Fast, low-cost language models could make real-time, note-based CDS practical, and current models are highly accurate. Prices and capabilities are changing rapidly, so continuous benchmarking and system updates are essential.

## 1 Background and Significance

Clinical decision support (CDS) systems, when implemented well, have been shown repeatedly to improve the quality and safety of care.[1–4] However, CDS is only as good as the data it runs on. Most CDS rules are written against coded, structured data in the electronic health record (EHR), such as problem lists, medication lists, laboratory results, and discrete fields for smoking status. When those data are missing or wrong, CDS fails to fire when it should (a false negative) or fires when it should not (a false positive). In prior work, we found that errors in the underlying data and in the logic that reads it are common causes of CDS malfunctions,[5–7] and we have worked to develop methods to infer missing structured data, such as problem list entries, from other parts of the record.[8–11]

Much of the information that CDS needs is not coded at all. It lives in free-text notes, where it is recorded as part of the clinical narrative.[12–14] Natural language processing (NLP) has long been used to recover this information,[15, 16] and identifying a patient’s smoking status is one of the canonical NLP extraction tasks. The 2006 i2b2 smoking challenge, for example, asked teams to classify patients as current, past, or never smokers from discharge summaries,[17, 18] and later systems extracted more detailed tobacco use information, including quantities and dates.[19–21] More recently, large language models (LLMs) have shown that they can extract clinical information from notes with little or no task-specific training,[22–26] including smoking status and smoking history.[27, 28] Our group has also used LLMs to improve CDS, for example, to suggest improvements to alert logic, to summarize alert override comments, and to identify documented barriers to statin therapy in notes,[29–31] and we have written about how generative AI should transform CDS.[32]

Lung cancer screening (LCS) is a good example of CDS that depends on information in notes. The United States Preventive Services Task Force (USPSTF) recommends annual screening with low-dose computed tomography for adults aged 50 to 80 years who have at least a 20 pack-year smoking history and who currently smoke or quit within the past 15 years.[33] The American Cancer Society (ACS) updated its guideline in 2023 to remove the years-since-quitting criterion,[34] and the Centers for Medicare & Medicaid Services uses criteria similar to those of the USPSTF.[35] Despite these recommendations, only 18.1% of eligible adults in the United States were up to date with screening in 2022.[36] CDS can help identify eligible patients and prompt shared decision-making,[37, 38] but every one of these criteria except age depends on smoking history. Structured smoking fields in the EHR usually record whether a patient smokes, but pack-years and quit dates are often missing, out of date, or inaccurate.[39–41] The information needed to determine eligibility is frequently recorded only in a progress note (for example, “former smoker, 1 ppd x 30 yrs, quit 2012”). In prior work at Vanderbilt University Medical Center (VUMC), we fine-tuned an NLP tool to extract quantitative smoking information from notes. Combining it with structured data identified 73.8% more patients eligible for LCS than structured data alone.[42]

LLMs could, in principle, fill these gaps for many CDS rules. In practice, we and others most often run LLMs in batch mode: notes are sent to a model offline (for example, overnight), and the extracted values are stored for later use.[43, 44] Batch processing is expensive and slow. A health system has millions of notes, and a general-purpose LLM takes seconds to read each one and charges for every token it reads and writes.[43] It would be preferable to run the model in real time, for example, by reading a patient’s recent notes when a primary care visit is opened, so that an LCS reminder can fire (or not) based on current information. With current general-purpose LLMs, this is usually too slow to fit within an EHR workflow and too expensive to run for every eligible patient.

One possible solution is a class of smaller, specialized models that trade the open-ended generality of chat-based LLMs for speed and cost. This class is broad. It includes encoder models such as BERT and its clinical variants, which are fast but usually need task-specific training data;[45–47] zero-shot classifiers built on natural language inference;[48] generalist extraction models such as GLiNER;[49] few-shot methods such as SetFit;[50] small generative models that can run on local hardware;[51] and techniques that constrain a generative model’s output to a fixed schema.[52] If a model of this type were cheap, fast, and accurate enough, it would enable many CDS use cases that are impractical today, such as checking notes for relevant history each time a rule is evaluated, rather than relying on data extracted the night before. To be used with patient data, such a model would also need to run locally or be hosted by a vendor under a business associate agreement (BAA) as required by the Health Insurance Portability and Accountability Act (HIPAA).[53, 54]

We thought that TypeSafe’s Jev model might be a useful example of this class. TypeSafe describes Jev as a “System One” model, a name taken from the fast, intuitive mode of thinking described by Kahneman.[55, 56] Jev reads natural language like an LLM, but it does not generate text. Instead, a developer sends it a *state* (text, or a JavaScript Object Notation [JSON] object) and a set of named questions, and Jev returns a typed answer for each question.[57, 58] There are three question types:

- **Choice**: select one option from a set that the developer defines (e.g., current, former, never, or unknown). The answer includes the selected option, a probability for each option, and a confidence value.
- **Score**: rate the state against ordered, descriptive levels. The answer includes a probability-weighted score, a probability for each level, and a confidence value.
- **Noul**: a yes/no question. The answer is the probability that the answer is yes.

The confidence value is a statistic computed from the shape of the probability distribution: a distribution concentrated on one option gives high confidence, and a flat distribution gives low confidence.[59] According to the vendor, Jev is trained with “reinforcement learning for calibrated decisions” so that its probabilities are calibrated, rather than being trained to produce text that people prefer.[60] Calibration here has its usual meaning: across many answers, outcomes assigned a probability of 0.8 should occur about 80% of the time.[61, 62] Because Jev cannot generate text, it cannot be used as a chatbot. It does not explain its answers, it cannot write out a number or a date that is not among the options it is given, and the vendor recommends keeping arithmetic and date calculations in the calling code.[63] For extraction tasks, the vendor recommends finding candidate values with simple code, such as regular expressions, and asking Jev to choose among them. Jev 1.13 accepts up to 64,000 tokens per request, and TypeSafe charges $0.042 per million input tokens, with no charge for output tokens.[64] By comparison, Anthropic’s Claude Haiku 4.5 and Claude Sonnet 5 cost $1 and $2 per million input tokens, respectively, and $5 and $10 per million output tokens.[65, 66]

To compare these models fairly, we needed a benchmark of clinical notes with known answers. The 2006 i2b2 smoking dataset would have been a natural choice, but the n2c2 portal, which distributes it, had marked its datasets as temporarily unavailable by September 2024,[67] and their data use agreement likely would not have allowed us to send the notes to commercial LLM services in any case (a concern that PhysioNet has also raised for its credentialed datasets).[68] Real notes from our own institution could not be shared with other researchers. We therefore built a synthetic benchmark, which can be shared openly.[69–72]

In this study, we developed a synthetic benchmark of 3,000 outpatient notes with known smoking status, pack-years, and quit dates, and we used it to compare a structured-judgment model (TypeSafe Jev 1.13) with four general-purpose LLMs (Claude Haiku 4.5, Claude Sonnet 5, GPT-6 Luna, and GPT-6 Sol). We measured accuracy, cost, and speed, and we assessed how each system’s errors would affect LCS eligibility decisions. We hypothesized that Jev would be substantially cheaper and faster than the LLMs, with similar accuracy on routine documentation, and that its accuracy would fall for documentation patterns that its question set did not anticipate.

## 2 Materials and Methods

Our study had two parts (Figure 1). First, we built a synthetic benchmark of three conditions with 1,000 notes each. We generated each synthetic patient’s smoking history and the corresponding reference labels in code, and only then produced a clinical note that conveyed that history. Second, we ran three systems on every note and scored both the extracted values and the LCS eligibility decisions that would follow from them.

**Figure 1:**
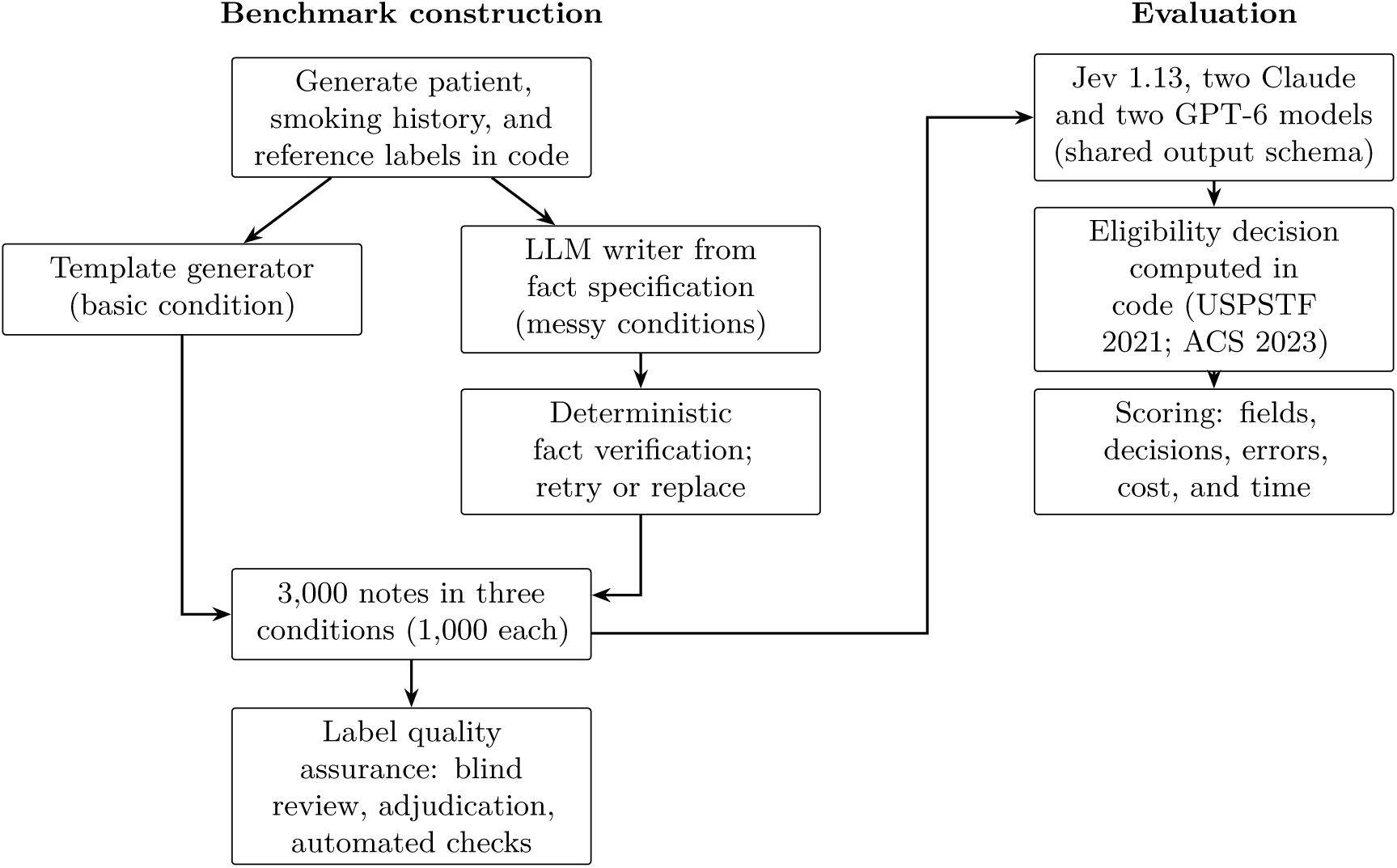
Study design. Reference labels were fixed before each note was generated. The same notes were given to every system, and each system’s extracted values were converted to a screening decision by the same code.

### 2.1 Benchmark construction

#### 2.1.1 Variables and reference labels

Each note in the benchmark was labeled with three variables. *Smoking status* was one of current, former, never, or unknown, and referred to tobacco cigarette smoking as of the visit date. Patients who were cutting down, had set a future quit date, or had relapsed after an earlier quit were current smokers; patients who had quit, however recently, were former smokers, even if they now used e-cigarettes or nicotine replacement; patients who had smoked fewer than 100 cigarettes were never smokers; and status was unknown when the note did not establish it. Cigars, pipes, cannabis, e-cigarettes, smokeless tobacco, secondhand smoke, and family members’ smoking did not count. *Pack-years* was packs per day multiplied by years smoked (one pack is 20 cigarettes). It was taken from a stated value when one was given, or computed when the note provided enough information (summing periods at different rates, using start and stop ages or years, excluding stated breaks, and using the midpoint of a stated range). It was 0 for never smokers and missing (null) when it could not be determined, for example, when a note gave only a rate (“smokes 1 ppd”) or only a duration (“x39y”). *Quit date* applied only to former smokers and was recorded at the precision the note supported (year, month, or day). Relative expressions such as “quit 10 years ago” or “quit at age 55” were resolved against the visit date and the patient’s age.

Because some notes are inherently imprecise, each reference label included an acceptable range. For example, “1 ppd since age 17” in a 60-year-old implies about 43 years of smoking, depending on the patient’s birthday, so we accepted 42 to 44 pack-years, and “quit 10 years ago” supports a quit year within one year of the visit year minus 10. Ranges were exact for stated values.

#### 2.1.2 Lung cancer screening eligibility

We used a simplified eligibility framework based only on age, pack-years, and, for former smokers, years since quitting. Under the USPSTF 2021 criteria, a patient was eligible if they were 50 to 80 years old, had at least 20 pack-years, and currently smoked or had quit 15 or fewer years before the visit.[33] We calculated years since quitting as the visit year minus the quit year. Age came from demographic data, as it would in an EHR, and all benchmark patients were 50 to 80 years old. We computed a three-level decision for every note: eligible, not eligible, or insufficient information (for example, a former smoker with at least 20 pack-years and no documented quit date). Some decisions could be made with partial information: a never smoker, or a former smoker who quit more than 15 years ago, is not eligible regardless of pack-years. When a reference range straddled a threshold (e.g., “15–25 pack-years”), we accepted either decision. We also computed decisions under the ACS 2023 guideline, which drops the years-since-quitting criterion.[34] The decision logic was implemented once, in code, and applied identically to the reference labels and to every system’s

#### 2.1.3 Basic condition

We wrote a Python program that generated 1,000 synthetic patients and one outpatient progress note for each. Patients were 50 to 80 years old (uniformly distributed), and visit dates fell between January 2023 and June 2026. For each patient, the program first sampled a smoking status (never 32%, former 35%, current 23%, unknown 10%) and, for smokers, a latent history: an age at starting (usually 15 to 20 years), a rate (most often one pack per day), and, for former smokers, a time since quitting drawn from a mixture that ranged from a few months to more than 40 years. It then chose how the note would document that history (Table 1) and computed the reference labels from that plan, so that the labels reflected what the note actually said rather than latent facts the note never stated. Relative quit dates (“N years ago” or “N months ago”) were used for about 15% of former smokers.

**Table 1:** How smoking histories were documented in the benchmark (examples paraphrased from generated notes).

| Element | Forms in the basic and messy conditions | Additional forms in the complex condition |
| --- | --- | --- |
| Pack-years | Stated (“40 PY”); packs per day × years; cigarettes per day × years; “since age 17”; “from age 20 to 50”; ranges (“1–2 ppd × 30 yrs”); EHR discrete fields (“Packs/day: 1.00; Years: 30.00”); insufficient (rate only, duration only, or vague) | Two or three periods at different rates; mixed packs and cigarettes; quit then restarted; start age plus quit calendar year; “since 1985”; “from 1978 until 2010” |
| Quit date | Year; month and year; full date; “N years ago”; “N months ago”; “at age 55”; tied to an event (“after his MI in 2016”); part of a decade (“mid-2000s”); multiple quit attempts; not documented | “3 years after her husband died in 2010”; “since his 60th birthday”; “when her grandson, now 7, was born”; event year in another section of the note |
| Distractors | Relapse after an earlier quit; future quit date; alcohol quit date; family members’ smoking; secondhand smoke; e-cigarettes, cannabis, cigars, or smokeless tobacco; “tried a few cigarettes”; a stale “never smoker” template field corrected in the narrative; copied-forward text from an earlier visit | Same |

The program rendered each note in one of six formats: an EHR-style note with discrete tobacco fields modeled on Epic Systems (Verona, Wisconsin, United States), a SOAP (subjective, objective, assessment, and plan) note, a terse note, a dictated narrative or consult letter, an assessment-first note, and a brief telehealth note. Each note included a chief complaint and history for one of 18 visit types (e.g., hypertension follow-up, annual examination, chest pain, or preoperative evaluation), comorbidities and consistent medications, vital signs, an examination, and an assessment and plan. Smoking information could appear in the history, the social history, a templated block, the problem list (e.g., “Nicotine dependence, cigarettes, uncomplicated (F17.210)”), or the assessment and plan, and was sometimes split across sections. Terse and assessment-first notes used heavy clinical abbreviations, and about 9% of notes contained one or two misspellings (e.g., “smoekr” or “qiut”). For patients with unknown status, the note had no social history, a social history that did not mention tobacco, “noncontributory”, “unable to obtain”, a declined question, or an EHR field reading “Unknown if ever smoked”.

#### 2.1.4 Messy and messy-with-complex-arithmetic conditions

Template-generated notes follow a limited set of phrasings, so we created two additional conditions in which an LLM wrote the notes. For each patient, code first generated a fact specification: demographics, visit type, note type (e.g., a nurse intake followed by a physician note, or a resident note with an attending attestation), known conditions, a target length (450 to 1,500 words), three to five “messiness” features to include, and a list of required facts and distractors. The messiness features included copied-forward text from an older visit, a templated social history with blank fields, long boilerplate, speech-recognition errors in non-smoking text, heavy abbreviations, an outdated problem list with ICD-10 codes, an addendum, direct patient quotes, and smoking information split across sections. Each required fact had machine-checkable values (for example, “The patient smoked 2 packs per day for 10 years, then cut back to 1 pack per day for 10 years before quitting” required the values 2, 10, 1, and 10). Code computed the reference labels from these facts.

Compared with the basic condition, these conditions enriched cases near the LCS thresholds. For 45% of smokers, the target pack-years were drawn from 12 to 28, and for 45% of former smokers, the years since quitting were drawn from 10 to 20. Years smoked were set to realistic spans (from the starting age to the visit or to the quit date), and rates were chosen to approach the target pack-years. The messy condition used documentation forms similar to the basic condition. The messy-with-complex-arithmetic condition (hereafter, the complex condition) used the additional forms in Table 1, which require multi-step arithmetic or date reasoning.

We used Claude Sonnet 5 (Anthropic, San Francisco, California, United States; effort setting “low”) to write each note. The instructions required the model to convey every fact exactly (in any wording, with numbers as digits or words), to add no other information about the patient’s own smoking, to avoid stating whether the patient was eligible for LCS or had a screening computed tomography ordered, and to return, for each fact, a verbatim quote from the note that conveyed it. Code then verified each note without using any model. It checked that each quote appeared verbatim in the note, that each quote contained the fact’s values (accepting number words, fractions such as “half a pack”, cigarette equivalents of pack rates, and two-digit years), that the note did not state a pack-year figure unless a fact gave one, that it made no statement about screening eligibility, and that it was at least 150 words long. Notes that failed were returned to the model with the specific problems, up to three attempts in total; specifications that still failed were replaced by the next specification from the generator. In the messy condition, the model made 1,343 calls: 696 notes passed on the first attempt, 286 on the second, and 18 on the third, and 7 specifications were replaced. In the complex condition, the corresponding numbers were 1,352 calls (plus one that failed with a server error); 682, 301, and 17 notes; and 6 replaced specifications. Writing and verification took about 20 and 21 minutes and cost $30.94 and $31.40, respectively. Box 1 shows excerpts from a representative note in each of the three conditions.

#### 2.1.5 Quality assurance of the reference labels

We checked the reference labels in three ways. First, an LLM-based reviewer (Claude Opus 5) that was blinded to the labels independently abstracted 60 notes from the two LLM-written conditions (and, in an earlier round, 60 notes from a preliminary version of the basic condition), and agreed with the reference on all three variables for every note. Second, after the evaluation runs (described below), we adjudicated every note for which any of the three systems disagreed with the reference on any variable or on the eligibility decision (465 notes: 51 basic, 60 messy, and 354 complex). Adjudication was performed by LLM-based review agents (Claude Opus 5), each of which read the note, derived the answer from the rules above, and only then compared it with the reference and with the anonymized outputs of the three systems. Third, we ran automated consistency checks on all 3,000 notes (for example, flagging former smokers whose note stated a quit time that the reference did not include, or never smokers whose note described smoking quantities).

The adjudication and error analysis identified a small number of systematic problems in the generators, all of which we corrected in code and then re-derived the labels without changing any note. The problems were: a quit age or year stated within the smoking history (“smoked 1 ppd from age 18 until she quit at age 45”) while the reference recorded the quit date as not documented; a stated break (“first quit in 2003, restarted in 2008”) that was not subtracted when the history was given as start and stop points; two quit statements in the same note that implied adjacent years; acceptable quit-year ranges that extended past the visit date; and event-based quit times that the note writer had hedged (“about 4 years after”), for which we accepted one year on either side (17 notes). When a stated duration might or might not include a break, we accepted both readings. In total, these corrections changed the reference labels for 84 of 3,000 notes (5 basic, 9 messy, and 70 complex). After correction, every one of the 465 adjudicated answers was consistent with the reference labels. One additional note became discordant only after the labels were corrected (a stated break that Haiku did not subtract); we reviewed it and confirmed the reference. All results in this paper use the corrected labels.

### 2.2 Evaluation

#### 2.2.1 Systems evaluated

Every system received only the note text and had to return the same output: smoking status (one of four values), pack-years (a number or null), and quit date (a string in YYYY, YYYY-MM, or YYYY-MM-DD format, or null). We defined this shared output as a single Pydantic data model in Python.

**TypeSafe Jev 1.13** (model identifier jev-1.13.0; TypeSafe AI) was called through its Python software development kit (typesafe-sdk version 0.7.0). Following the vendor’s guidance,[58, 63] we sent each note as the state in a single request with about 34 Choice questions, and all arithmetic and date calculations were done in our code. The questions asked for the smoking status; how pack-years were documented (stated, a stated range, rate and years, rate from a start point, rate from a start point to a stop point, two or three periods at different rates, other or unclear, or not computable); the stated pack-year number (the options were the numbers, including number words, that a regular expression found in the note); rates in packs or cigarettes per day; years smoked; start and stop ages and years; the length of any break; the rate and duration of up to three periods; how the quit date was documented; the quit year, month, day, years or months before the visit, or decade; the patient’s age; and the visit date (the options were the dates found in the note). Each question included an option for “none”, and the two routing questions included an “other or unclear” option, which our code treated as not determinable. Our code then assembled the answers into the shared output, for example, by adding the pack-years of each period or subtracting the stated number of years from the visit year.

##### Box 1: Excerpts from a representative note in each condition, with the reference labels. The note text is reproduced verbatim, including abbreviations and errors; omitted text is marked […]. All notes are synthetic, and the names, dates, and identifiers in them are fabricated.

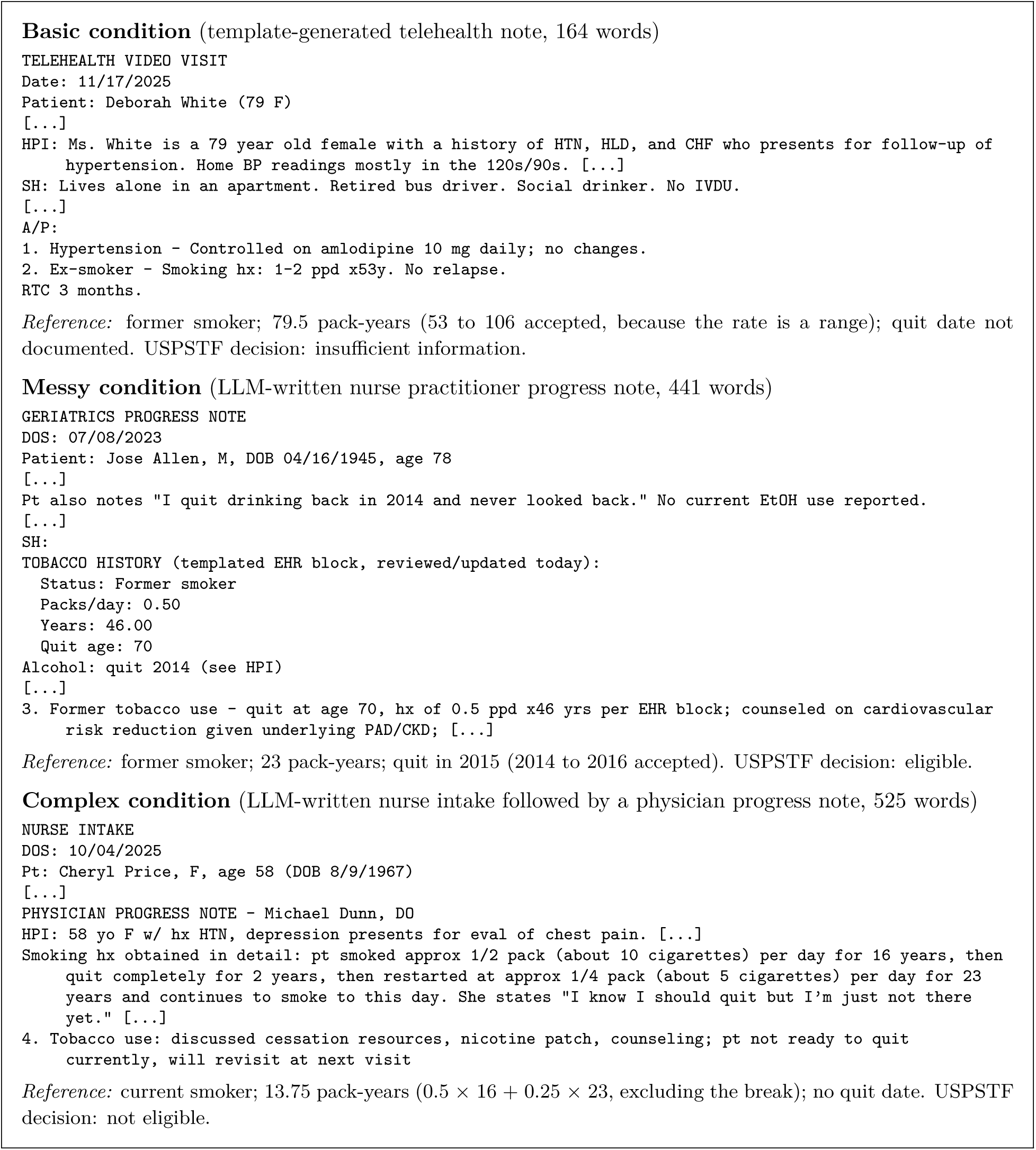

**Claude Haiku 4.5** (claude-haiku-4-5-20251001) and **Claude Sonnet 5** (claude-sonnet-5) were called through the Anthropic Python software development kit (version 1.7.0) with structured outputs, which constrain the model’s response to a JSON schema.[73] The system prompt contained the variable definitions and rules described above. The schema asked the model first for supporting evidence (the visit date, the patient’s age, verbatim quotes about the patient’s smoking, any stated pack-year figure, each smoking period with its rate and duration, the basis for the pack-year value, and a quote giving the quit timing) and then for the three shared output fields, which were the only fields we scored. We ran Sonnet 5 with its effort setting at “low”, a setting that controls how much reasoning the model does before answering, and we ran Haiku 4.5 with its default settings (no extended thinking). Both used a maximum of 16,000 output tokens. Because the system prompt and the schema are identical for every note, we marked them for prompt caching, which bills a repeated prefix at a tenth of the input price; this applied to Sonnet 5 but not to Haiku 4.5, whose minimum cacheable prefix of 4,096 tokens is longer than our prompt and schema.[74] The two OpenAI models cache repeated prefixes automatically.

**GPT-6 Sol** (gpt-6-sol) and **GPT-6 Luna** (gpt-6-luna) were released by OpenAI on September 22, 2026, three days after the runs described above, and we added them to the comparison the following day.[75] They were called through the OpenAI Python software development kit (version 3.19.0) with structured outputs, using the same system prompt, the same schema, and the same scoring code as the Claude runs, with no changes of any kind. Both are reasoning models whose reasoning effort can be set from none to max. We ran Sol at “low”, matching Sonnet 5, and Luna with reasoning off, matching Haiku 4.5, and both used a maximum of 16,000 output tokens. Sol is OpenAI’s mid-size model and Luna its high-volume model; their knowledge cutoffs (April 20 and May 18, 2026) precede the publication of this benchmark, so the notes could not have been in their training data.

We developed the Jev question set and the Claude prompt on a separate set of 40 notes generated with a different random seed. On that set, we compared Haiku 4.5 and Sonnet 5 (effort “low”) to choose a fast, accurate, and cost-effective configuration, and we evaluated both. We also ran a preliminary comparison with a shorter prompt and a smaller TypeSafe question set on the basic condition; that comparison informed the final design, and we do not detail it here. Before we designed or generated the messy and complex conditions, we froze the prompts, schemas, question sets, and scoring code and recorded their SHA-256 hashes, so that neither system could be tuned to the new notes.

#### 2.2.2 Execution and measurement

We ran all systems from Python 3.12 using asynchronous clients with 16 concurrent requests per system. For each condition, Jev and Sonnet ran at the same time; Haiku ran afterwards. The Jev and Claude runs took place on September 19, 2026, and the two OpenAI runs on September 23, 2026 (Coordinated Universal Time), in the same way and at the same concurrency. We used the software development kits’ automatic retries for rate limits and transient errors. One Haiku response in the messy condition returned a quit date that did not match the required format (“2010s”) and was scored as incorrect on all variables. We recorded the wall-clock time for each run of 1,000 notes, the latency of each request, and the input and output tokens reported by each application programming interface (API), and we calculated cost from the tokens and each vendor’s list price (Jev: $0.042 per million input tokens and no charge for output tokens; Haiku 4.5: $1 and $5 per million input and output tokens; Sonnet 5: $2 and $10; Sol: $2 and $10; Luna: $0.10 and $0.50).[64, 65, 76] We recorded the model version returned by each API call.

#### 2.2.3 Statistical analysis

We scored smoking status as correct if it matched the reference exactly, pack-years as correct if they fell within the acceptable range (with a tolerance of 0.5 pack-years for rounding) or were null when the reference was null, and quit date as correct if its year fell within the acceptable range or it was null when the reference was null. An eligibility decision was correct if it was one of the decisions allowed by the reference. To characterize errors in terms that matter for CDS, we treated a decision of eligible as a screening flag (i.e., the patient would be flagged for LCS by CDS) and calculated true positives, false positives (patients flagged who were not eligible), false negatives (eligible patients who were not flagged, whether the system decided they were not eligible or that there was insufficient information), sensitivity, specificity, and positive predictive value (PPV). We calculated 95% confidence intervals for proportions using the Wilson method and compared paired decision accuracy between systems with an exact McNemar test. We compared error counts between conditions with Fisher’s exact test.

#### 2.2.4 Projection to health-system scale

To estimate what it would cost to process notes at the scale of a large academic medical center, we used the VUMC Synthetic Derivative (SD), a de-identified copy of the VUMC EHR that currently contains more than 3.9 million patient records[77, 78] and that contained more than 200 million notes for more than 3.4 million patients as of December 2021.[79, 80] We multiplied the measured cost per note and throughput of each system by 200 million notes. We also estimated the cost of a real-time approach limited to the patients who could be affected by an LCS rule: the 102,475 patients aged 50 to 80 years with a primary care encounter at VUMC between 2019 and 2022 in our prior study,[42] with 50 notes per patient.

### 2.3 Ethics

This study did not involve human subjects, human samples, or human data. All patients and notes are synthetic, so institutional review board review was not required.

## 3 Results

### 3.1 Benchmark characteristics

Table 2 shows the composition of the three conditions. The LLM-written notes were more than twice as long as the template-generated notes (median 559 to 567 words vs 242 words). By design, the messy and complex conditions contained more smokers and more patients near the eligibility thresholds: 178 and 206 smokers had reference pack-years between 15 and 25, compared with 64 in the basic condition. In the basic condition, 250 notes (25.0%) documented a patient who was eligible for LCS under the USPSTF criteria, 495 (49.5%) a patient who was not eligible, and 249 (24.9%) a patient whose eligibility could not be determined from the note; 6 notes (0.6%) were borderline and accepted two decisions.

**Table 2:** Composition of the benchmark by condition. Eligibility is under the USPSTF 2021 criteria; borderline notes had a reference range that straddled a threshold, so two decisions were accepted.

|  | Basic | Messy | Complex |
| --- | --- | --- | --- |
| Notes | 1,000 | 1,000 | 1,000 |
| Words per note, median (IQR) | 242 (194–296) | 567 (448–698) | 559 (444–686) |
| *Smoking status* |  |  |  |
| Current | 226 | 336 | 314 |
| Former | 355 | 406 | 410 |
| Never | 324 | 180 | 199 |
| Unknown | 95 | 78 | 77 |
| Smokers with known pack-years, n/N (%) | 409/581 (70.4) | 611/742 (82.3) | 645/724 (89.1) |
| Former smokers with a quit date, n/N (%) | 307/355 (86.5) | 357/406 (87.9) | 381/410 (92.9) |
| Smokers with 15 to 25 pack-years | 64 | 178 | 206 |
| Former smokers who quit 12 to 18 years earlier | 70 | 118 | 111 |
| *Reference eligibility decision* |  |  |  |
| Eligible | 250 | 290 | 313 |
| Not eligible | 495 | 482 | 513 |
| Insufficient information | 249 | 208 | 154 |
| Borderline | 6 | 20 | 20 |

### 3.2 Extraction and eligibility accuracy

Table 3 and Figure 2 show the accuracy of each system. In the basic condition, Jev made the correct USPSTF eligibility decision for 991 of 1,000 notes (99.1%), compared with 997 (99.7%) for Luna, 978 (97.8%) for Haiku, and 1,000 (100.0%) for both Sol and Sonnet. Jev was less accurate than Sol and Sonnet (P = .004 for each), more accurate than Haiku (P = .015), and did not differ significantly from Luna (P = .15). In the messy condition, Jev was correct for 999 notes (99.9%),

**Figure 2:**
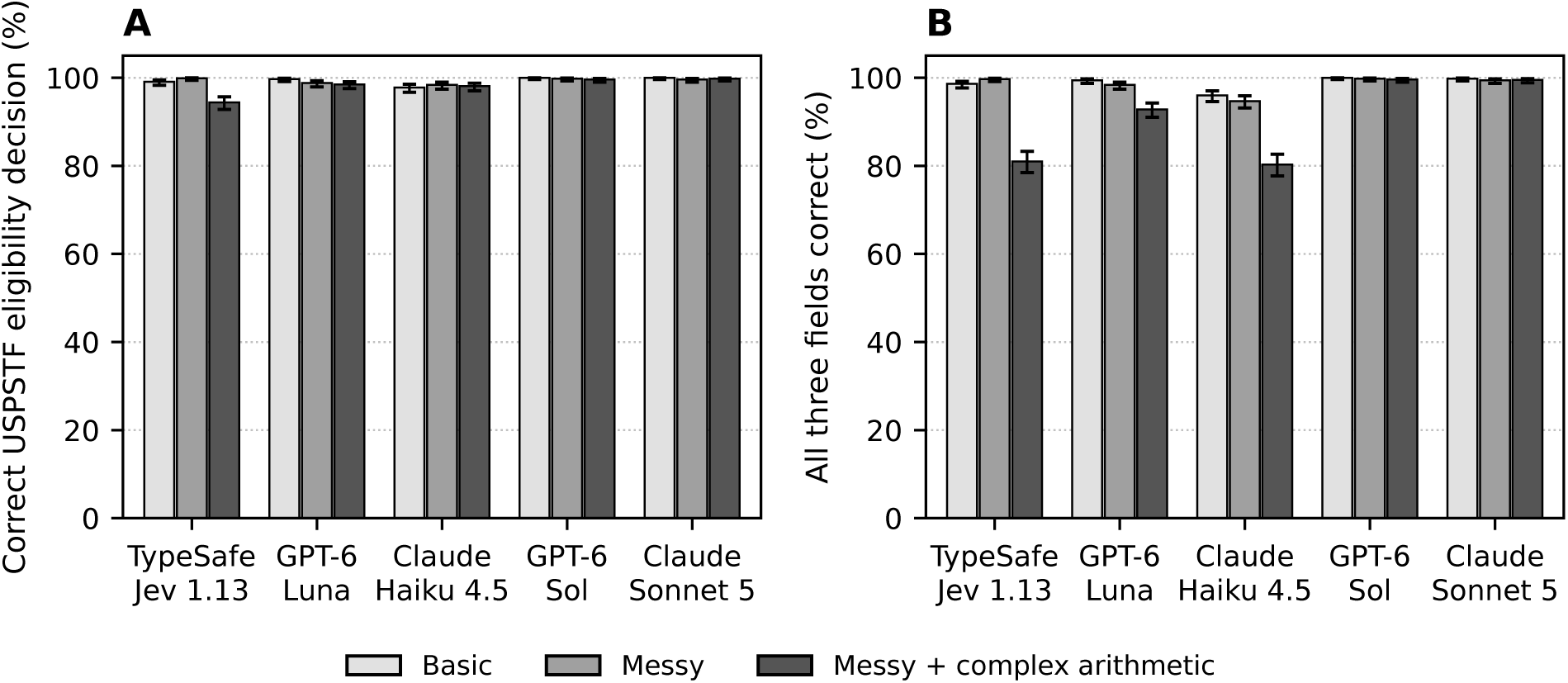
Accuracy by system and condition. (A) Correct USPSTF 2021 eligibility decisions. (B) Notes for which smoking status, pack-years, and quit date were all correct. Error bars show 95% confidence intervals.

**Table 3:**
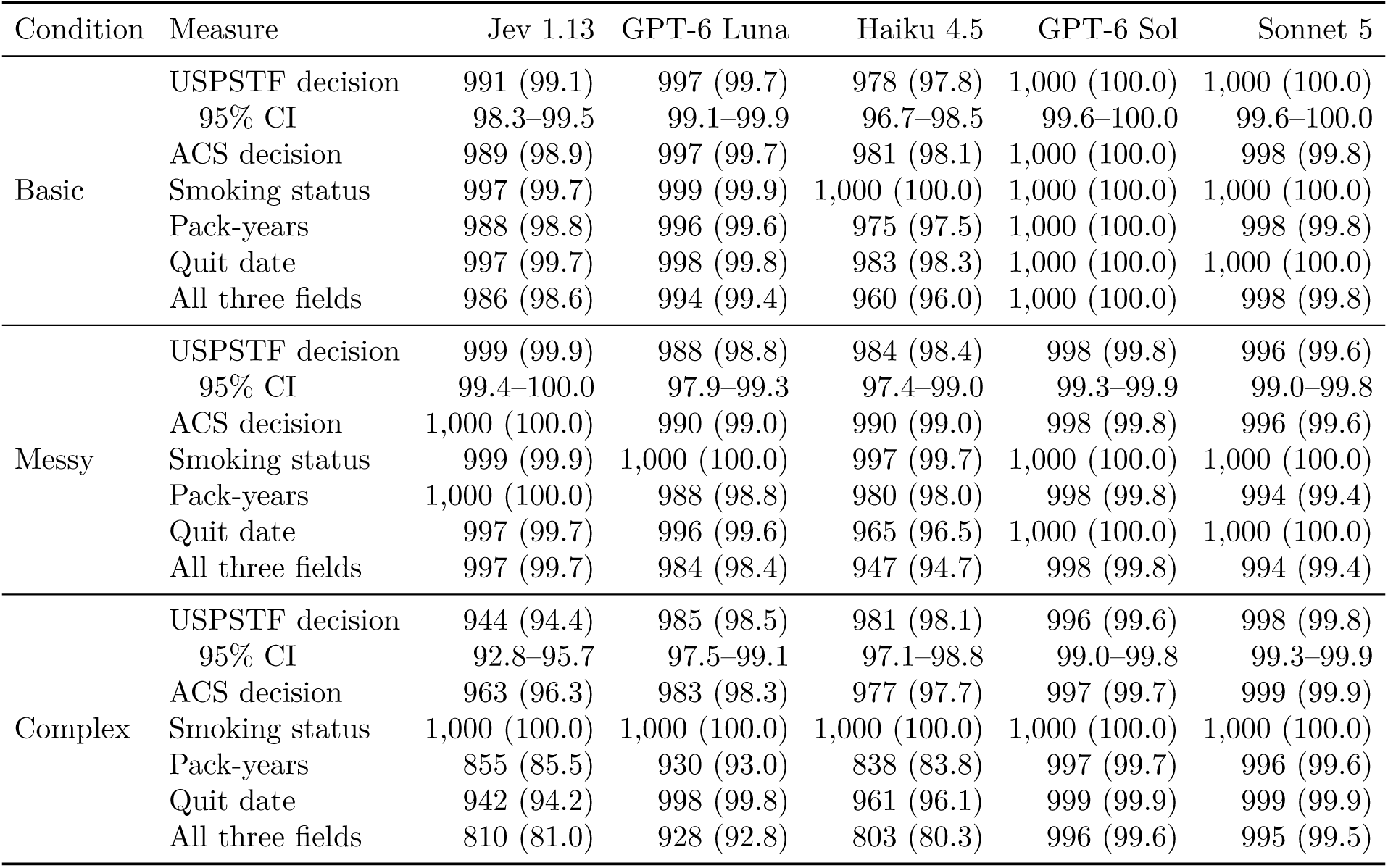
Accuracy by condition and system, n/1,000 (%). The 95% confidence interval (CI) is shown for the USPSTF eligibility decision.

Luna for 988 (98.8%), Haiku for 984 (98.4%), Sol for 998 (99.8%), and Sonnet for 996 (99.6%); Jev, Sol, and Sonnet did not differ from one another (P *≥* .38), and each was more accurate than Haiku (P *≤* .012). In the complex condition, Jev was the least accurate system, at 944 notes (94.4%), compared with 985 (98.5%) for Luna, 981 (98.1%) for Haiku, 996 (99.6%) for Sol, and 998 (99.8%) for Sonnet (P < .001 for Jev vs each). Sol and Sonnet did not differ in any condition (P *≥* .62). The results under the ACS 2023 guideline were similar.

All five systems identified smoking status almost perfectly, with no more than 3 errors per 1,000 notes for any system in any condition. The differences were in pack-years and quit dates. In the complex condition, Jev extracted pack-years correctly for 855 notes (85.5%) and quit dates correctly for 942 (94.2%); the corresponding results were 930 (93.0%) and 998 (99.8%) for Luna, 838 (83.8%) and 961 (96.1%) for Haiku, 997 (99.7%) and 999 (99.9%) for Sol, and 996 (99.6%) and 999 (99.9%) for Sonnet. Many field-level errors did not change the eligibility decision (for example, an error in pack-years for a patient with more than 60 pack-years, or an error in the quit date for a patient whose smoking history was well below 20 pack-years). As a result, eligibility decisions were more accurate than the fields from which they were derived.

### 3.3 Impact on lung cancer screening eligibility

Table 4 and Figure 3 show how each system’s errors would have affected a simple CDS rule that flags patients who are eligible for screening. In the basic and messy conditions, Jev produced 7 and 1 incorrect flags, respectively. In the complex condition, Jev flagged 27 patients who were not eligible (false positives) and failed to flag 14 eligible patients (false negatives), for a sensitivity of 0.957 and a PPV of 0.919. Twelve of Jev’s 14 false negatives were decisions that there was insufficient information. In a CDS context, these could prompt a clinician to review the smoking history rather than silently skipping the patient. Only 2 were decisions that an eligible patient was not eligible. In the complex condition, Haiku produced fewer false positives than Jev (4 vs 27) and a similar number of false negatives (13 vs 14), but all 13 of Haiku’s false negatives were decisions that an eligible patient was not eligible. Across the three conditions, Haiku failed to flag 30 eligible patients, compared with 16 for Jev. Luna produced 2, 4, and 10 incorrect flags in the three conditions, Sol produced 0, 0, and 3, and Sonnet produced 0, 2, and 0.

**Figure 3:**
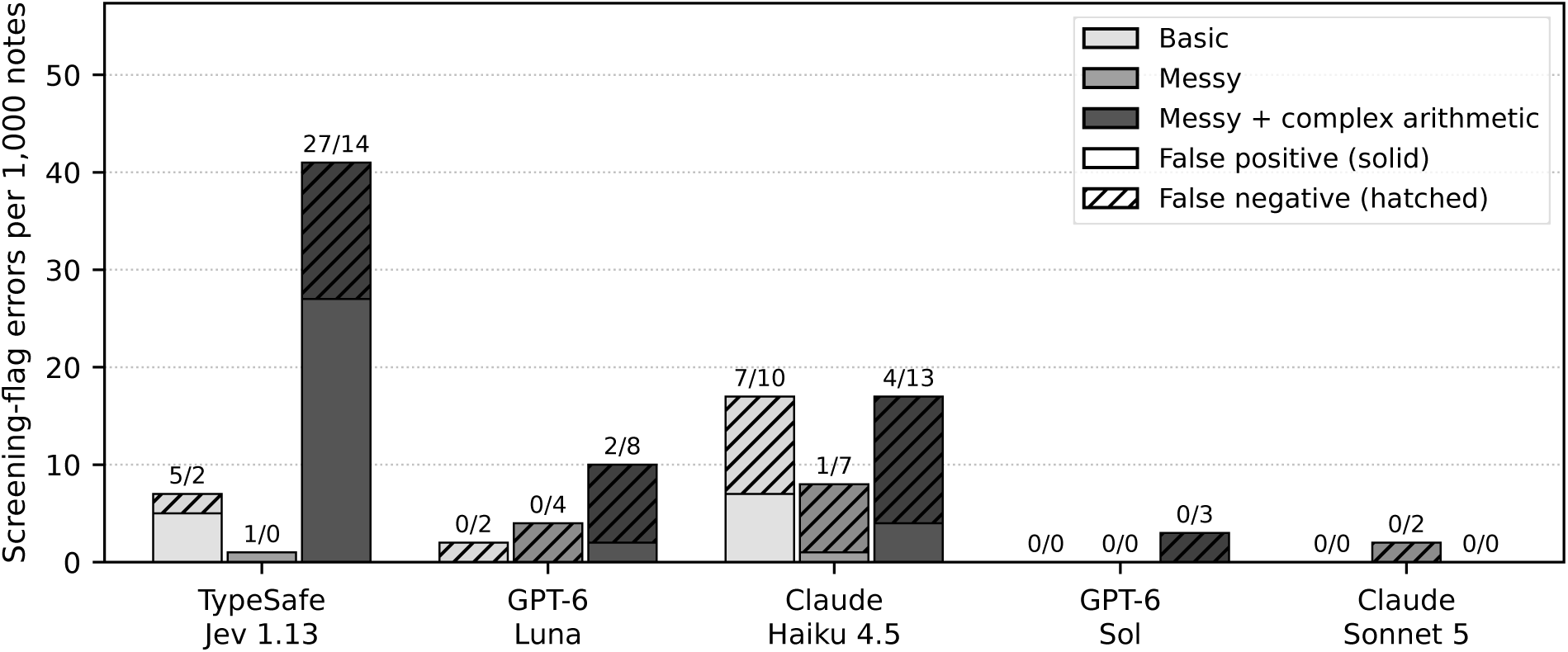
Incorrect screening flags per 1,000 notes under the USPSTF 2021 criteria. Solid segments are false positives (patients flagged who were not eligible), and hatched segments are false negatives (eligible patients who were not flagged). Labels show false positives/false negatives.

**Table 4:** Screening flags produced by each system under both guidelines. A patient was flagged when the system’s extracted values made them eligible. False negatives include eligible patients classified as not eligible and those classified as having insufficient information (in parentheses: not eligible / insufficient information). PPV, positive predictive value.

| Condition | System | TP | FP | FN | TN | Sensitivity | Specificity | PPV |
| --- | --- | --- | --- | --- | --- | --- | --- | --- |
| *USPSTF 2021* |  |  |  |  |  |  |  |  |
| Basic | Jev 1.13 | 250 | 5 | 2 (0/2) | 743 | 0.992 | 0.993 | 0.980 |
|  | GPT-6 Luna | 250 | 0 | 2 (0/2) | 748 | 0.992 | 1.000 | 1.000 |
|  | Haiku 4.5 | 242 | 7 | 10 (10/0) | 741 | 0.960 | 0.991 | 0.972 |
|  | GPT-6 Sol | 252 | 0 | 0 | 748 | 1.000 | 1.000 | 1.000 |
|  | Sonnet 5 | 252 | 0 | 0 | 748 | 1.000 | 1.000 | 1.000 |
| Messy | Jev 1.13 | 301 | 1 | 0 | 698 | 1.000 | 0.999 | 0.997 |
|  | GPT-6 Luna | 297 | 0 | 4 (1/3) | 699 | 0.987 | 1.000 | 1.000 |
|  | Haiku 4.5 | 292 | 1 | 7 (5/2) | 700 | 0.977 | 0.999 | 0.997 |
|  | GPT-6 Sol | 301 | 0 | 0 | 699 | 1.000 | 1.000 | 1.000 |
|  | Sonnet 5 | 299 | 0 | 2 (0/2) | 699 | 0.993 | 1.000 | 1.000 |
| Complex | Jev 1.13 | 308 | 27 | 14 (2/12) | 651 | 0.957 | 0.960 | 0.919 |
|  | GPT-6 Luna | 313 | 2 | 8 (4/4) | 677 | 0.975 | 0.997 | 0.994 |
|  | Haiku 4.5 | 306 | 4 | 13 (13/0) | 677 | 0.959 | 0.994 | 0.987 |
|  | GPT-6 Sol | 317 | 0 | 3 (0/3) | 680 | 0.991 | 1.000 | 1.000 |
|  | Sonnet 5 | 321 | 0 | 0 | 679 | 1.000 | 1.000 | 1.000 |
| *ACS 2023* |  |  |  |  |  |  |  |  |
| Basic | Jev 1.13 | 315 | 6 | 2 (0/2) | 677 | 0.994 | 0.991 | 0.981 |
|  | GPT-6 Luna | 315 | 0 | 2 (0/2) | 683 | 0.994 | 1.000 | 1.000 |
|  | Haiku 4.5 | 309 | 10 | 8 (8/0) | 673 | 0.975 | 0.985 | 0.969 |
|  | GPT-6 Sol | 317 | 0 | 0 | 683 | 1.000 | 1.000 | 1.000 |
|  | Sonnet 5 | 317 | 1 | 0 | 682 | 1.000 | 0.999 | 0.997 |
| Messy | Jev 1.13 | 410 | 0 | 0 | 590 | 1.000 | 1.000 | 1.000 |
|  | GPT-6 Luna | 407 | 0 | 3 (0/3) | 590 | 0.993 | 1.000 | 1.000 |
|  | Haiku 4.5 | 407 | 2 | 2 (0/2) | 589 | 0.995 | 0.997 | 0.995 |
|  | GPT-6 Sol | 410 | 0 | 0 | 590 | 1.000 | 1.000 | 1.000 |
|  | Sonnet 5 | 408 | 0 | 2 (0/2) | 590 | 0.995 | 1.000 | 1.000 |
| Complex | Jev 1.13 | 423 | 33 | 3 (3/0) | 541 | 0.993 | 0.943 | 0.928 |
|  | GPT-6 Luna | 416 | 4 | 8 (4/4) | 572 | 0.981 | 0.993 | 0.990 |
|  | Haiku 4.5 | 414 | 9 | 11 (11/0) | 566 | 0.974 | 0.984 | 0.979 |
|  | GPT-6 Sol | 421 | 0 | 2 (0/2) | 577 | 0.995 | 1.000 | 1.000 |
|  | Sonnet 5 | 425 | 0 | 0 | 575 | 1.000 | 1.000 | 1.000 |

The choice of guideline changed which extraction errors mattered (Table 4). Under the ACS 2023 criteria, which do not use the quit date, Jev’s sensitivity in the complex condition rose from 0.957 to 0.993, because most of its false negatives under the USPSTF criteria came from a missing or wrong quit date rather than from pack-years. At the same time its specificity fell from 0.960 to 0.943 and its false positives rose from 27 to 33, since pack-year errors alone then determine eligibility. The other systems moved much less, and the ordering of the five systems was the same under both guidelines.

### 3.4 Errors by documentation pattern

Errors were concentrated in a small number of documentation patterns (Table 5). There are several explanations for Jev’s errors in the complex condition. First, when a patient quit and later restarted smoking, Jev answered the questions about each smoking period independently and usually reported a spurious third period, which our code then added. This overstated pack-years in 97 of 100 such notes; in 81 of them, the spurious period was a copy of the second. TypeSafe’s documentation warns about this kind of problem. It states that “Jev is not a calculator,” that it “does not count reliably,” and that it “struggles with tasks that require numeric precision,” and it notes that asking “which of two dates comes first, how far apart they are, or whether one falls inside a window is unreliable.”[63] We kept all arithmetic and comparisons in our code, as the vendor recommends, but deciding how many periods a note described was, in effect, a counting task that we left to the model. Second, some patterns were not anticipated by the question set. For example, Jev had no way to express “she quit when her grandson, now 7, was born”, and it answered only 8 of 48 of these quit dates correctly. Similarly, “quit 3 years after her husband died in 2010” requires adding 3 to 2010, which Jev’s options did not support directly, and Jev answered 49 of 65 correctly. Third, Jev handled patterns that fit its question set well, including three periods at different rates (84 of 84), “since 1985” (40 of 40), and quit dates tied to a birthday or an age (52 of 52 and 43 of 43).

**Table 5:**
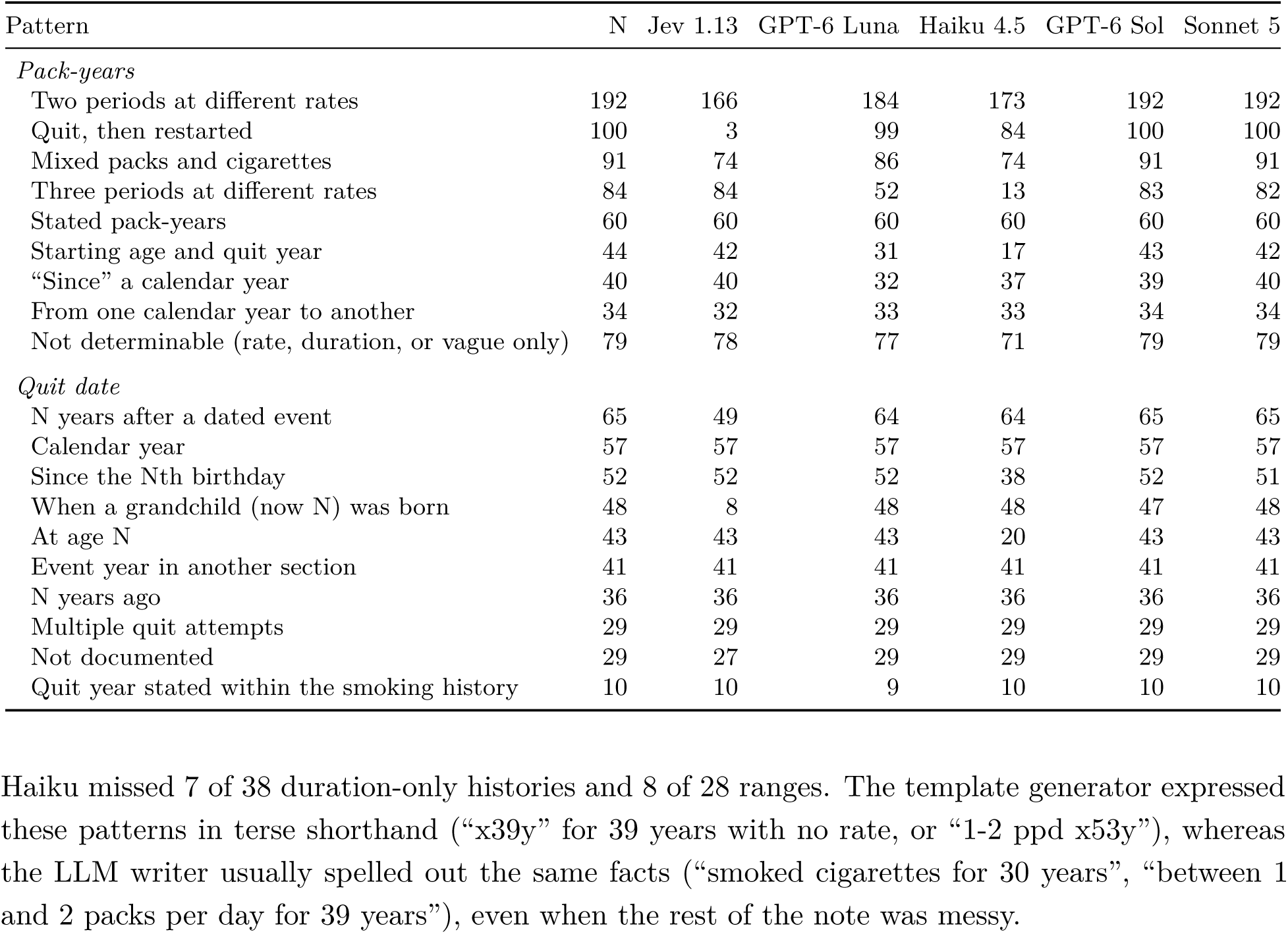
Correct extraction by documentation pattern for current and former smokers in the complex condition, n/N. Pack-year patterns are scored on pack-years, and quit-date patterns on quit date.

Haiku’s errors were mostly arithmetic. It computed pack-years correctly for only 13 of 84 notes with three periods at different rates and 17 of 44 notes that combined a starting age with a quit year, and it resolved “quit at age 55” correctly in 20 of 43 notes; in 21 of its 23 errors, it returned a year at least 14 years too early. Sonnet and Sol made few errors of any kind: five in total across every pattern for Sonnet, and four for Sol. Luna failed on the same arithmetic patterns as Haiku, but less often: it computed pack-years correctly for 52 of 84 notes with three periods at different rates and 31 of 44 that combined a starting age with a quit year. Unlike Jev, it handled quit-and-restart histories (99 of 100) and quit dates defined by a grandchild’s current age (48 of 48).

Jev and Haiku made more errors in the basic condition than in the messy condition (for Jev, 9 vs 1 incorrect decisions, P = .02; for Haiku, 22 vs 16, P = .41). The same documentation patterns were easier to read in the LLM-written notes. For example, in the messy condition, Jev made the correct decision for all 52 notes that gave a range and all 55 that gave only a duration, and Haiku for 52 of 52 and 53 of 55. In the basic condition, Jev missed 5 of 38 duration-only histories, and Haiku missed 7 of 38 duration-only histories and 8 of 28 ranges. The template generator expressed these patterns in terse shorthand (“x39y” for 39 years with no rate, or “1-2 ppd x53y”), whereas the LLM writer usually spelled out the same facts (“smoked cigarettes for 30 years”, “between 1 and 2 packs per day for 39 years”), even when the rest of the note was messy.

### 3.5 Cost and speed

Table 6 and Figure 4 show the cost and speed of each system. Luna was the least expensive system, at $0.12 to $0.21 per 1,000 notes, followed by Jev at $0.61 to $0.64, Sol at $2.44 to $4.22, Haiku at $3.37 to $4.43, and Sonnet at $3.76 to $6.61. Luna cost three to five times less than Jev and about 30 times less than Sonnet. For the models that support it we cached the system prompt and the output schema, which are the same for every note; with caching off, Sonnet cost $8.54 to $11.42 per 1,000 notes, and Haiku’s prompt was too short to be cached at all. Most of Jev’s roughly 15,000 input tokens per note were the answer options for its questions, not the note itself, so its cost was nearly constant across conditions. Jev had the lowest median latency by a wide margin (0.45 to 1.21 seconds per note, compared with 1.71 to 2.41 seconds for Haiku, 2.29 to 3.40 for Luna, 2.46 to 3.50 for Sonnet, and 2.51 to 3.39 for Sol) and the highest throughput; it was the only system that answered in under a second in any condition. Jev’s latency varied between runs with similar workloads (for example, 0.45 seconds in the complex condition and 1.21 seconds in the messy condition), which likely reflects load on the vendor’s service rather than the notes themselves.

**Figure 4:**
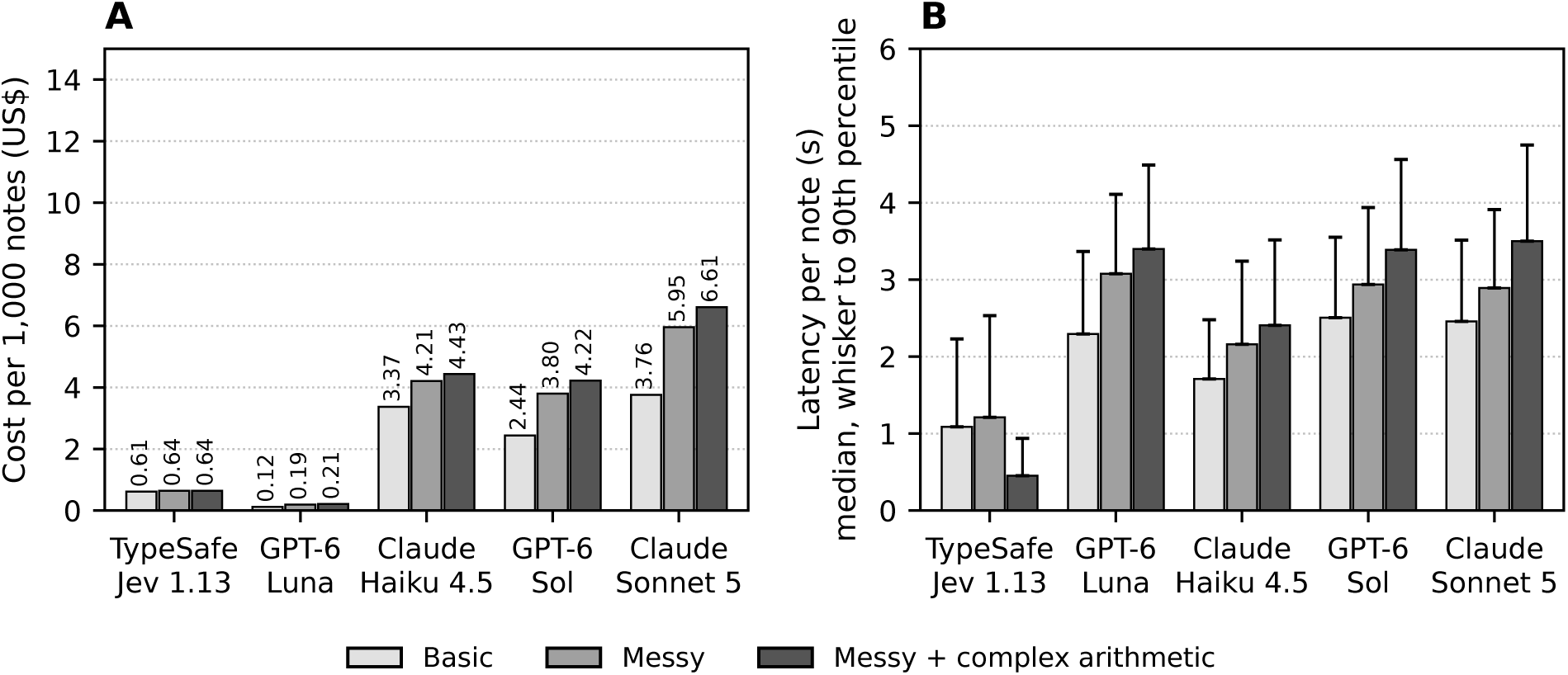
Cost and speed. (A) Cost per 1,000 notes. (B) Median latency per note; whiskers extend to the 90th percentile.

**Table 6:** Cost and speed for 1,000 notes, with 16 concurrent requests per system. Cost was calculated from the tokens reported by each API and each vendor’s list price.

| Condition | System | Cost (US\$) | Wall-clock (s) | Latency per note (s) | | Input tokens per note |
| --- | --- | --- | --- | --- | --- | --- |
|  |  |  |  | Median | 90th pct. |  |
| Basic | Jev 1.13 | 0.61 | 85 | 1.09 | 2.23 | 14,618 |
|  | GPT-6 Luna | 0.12 | 174 | 2.29 | 3.37 | 1,801 |
|  | Haiku 4.5 | 3.37 | 119 | 1.71 | 2.48 | 2,720 |
|  | GPT-6 Sol | 2.44 | 186 | 2.51 | 3.55 | 1,801 |
|  | Sonnet 5 | 3.76 | 170 | 2.46 | 3.51 | 3,463 |
| Messy | Jev 1.13 | 0.64 | 92 | 1.21 | 2.53 | 15,285 |
|  | GPT-6 Luna | 0.19 | 206 | 3.08 | 4.11 | 2,303 |
|  | Haiku 4.5 | 4.21 | 148 | 2.16 | 3.24 | 3,299 |
|  | GPT-6 Sol | 3.80 | 202 | 2.94 | 3.94 | 2,303 |
|  | Sonnet 5 | 5.95 | 194 | 2.89 | 3.91 | 4,301 |
| Complex | Jev 1.13 | 0.64 | 39 | 0.45 | 0.94 | 15,272 |
|  | GPT-6 Luna | 0.21 | 217 | 3.40 | 4.49 | 2,290 |
|  | Haiku 4.5 | 4.43 | 160 | 2.41 | 3.52 | 3,288 |
|  | GPT-6 Sol | 4.22 | 225 | 3.39 | 4.56 | 2,290 |
|  | Sonnet 5 | 6.61 | 224 | 3.50 | 4.75 | 4,279 |

At these rates, processing the more than 200 million notes in the VUMC SD once would cost about $24,000 to $43,000 with Luna, $123,000 to $128,000 with Jev, $488,000 to $844,000 with Sol, $674,000 to $887,000 with Haiku, and $752,000 to $1.3 million with Sonnet. At the throughput we observed with 16 concurrent requests, it would take about 91 to 213 days with Jev, 276 to 371 days with Haiku, 393 to 519 days with Sonnet, 403 to 503 days with Luna, and 430 to 520 days with Sol, although higher concurrency, where the vendor’s rate limits allow it, would shorten these times. By contrast, reading 50 notes for each of the 102,475 primary care patients aged 50 to 80 years in our prior study would cost about $610 to $1,100 with Luna, $3,100 to $3,300 with Jev, $12,500 to $21,600 with Sol, $17,000 to $23,000 with Haiku, and $19,000 to $34,000 with Sonnet.

The benchmark itself was inexpensive to build. Generating and verifying the 2,000 LLM-written notes cost $62.34 and took about 41 minutes, and the template-generated notes cost nothing and took seconds to produce. Running all five systems on all 3,000 notes cost $41.21 in total at list prices.

## 4 Discussion

A structured-judgment model, TypeSafe Jev 1.13, extracted smoking histories from clinical notes with accuracy similar to general-purpose LLMs on routine documentation and with lower latency than any of them. Jev made the correct LCS eligibility decision for 99.1% of template-generated notes and 99.9% of messy LLM-written notes. However, its accuracy fell to 94.4% when notes required complex arithmetic or used phrasings that its question set did not anticipate, while Claude Sonnet 5 and GPT-6 Sol remained above 99% in every condition, and GPT-6 Luna, the cheapest system we tested, was correct for 98.5% of the complex notes at a third of Jev’s cost per note.

The impact of an extraction error depends on which guideline the rule implements. Under the USPSTF criteria, the quit date decides eligibility for every former smoker, and a missing or wrong quit date accounted for 12 of Jev’s 14 false negatives in the complex condition. The ACS criteria drop the years-since-quitting requirement, so those errors cost almost nothing there and Jev’s sensitivity rose from 0.957 to 0.993. Pack-year errors matter under both guidelines, and with the quit date out of the calculation they accounted for all 33 of Jev’s false positives under ACS, up from 27 under USPSTF. To accurately characterize the clinical implications of potential extraction errors, tools like these should be evaluated against the rules they will actually feed.

This comparison changed twice while we were writing it. We got access to Jev on a Friday, built the benchmark and ran it against the two Claude models that night (September 19), and wrote the first version of this paper with a clear conclusion: a model that makes typed judgments rather than text was almost as accurate as a frontier LLM on ordinary notes, cost a fraction as much, and was the only system fast enough for real-time use. Three days later, on September 22, 2026, OpenAI released GPT-6 Sol and GPT-6 Luna. We ran both the next day, with the prompt and schema unchanged. Sol was as accurate as Sonnet on eligibility decisions in every condition (P *≥* .62) at about two-thirds of the price, and Luna, the high-volume model, cost meaningfully less per note than Jev while making fewer eligibility errors than Jev on the complex notes. The conclusion we had written four days earlier, that a structured-judgment model wins on cost, was no longer true. The pace is dizzying. For anyone building CDS on these models, the practical lesson is that the choice of model is temporary. The benchmark, the prompts, the scoring code, and the eligibility logic are still durable, and should be built modularly so that a new model can be dropped into them in an afternoon, as we did here.

Overall, we believe these results are encouraging. For most real-world CDS, a model does not need to be perfect. It needs to be accurate enough, fast enough, and cheap enough to be used where the decision is made. The most accurate Claude model in our study, Sonnet, may be too expensive and too slow for many of the uses we would most like to support. At the prices and throughput we observed, a single pass over the more than 200 million notes in the VUMC SD would cost about $752,000 to $1.3 million with Sonnet and would take 393 to 519 days at the concurrency we used, compared with about $123,000 to $128,000 with Jev and $24,000 to $43,000 with Luna. These estimates are likely conservative for the LLMs, whose cost scales with the length of each note: our synthetic notes had a median of 242 to 567 words, and many real notes are longer. They also cover only one use case. A health system that wanted to extract five or ten different variables for different CDS rules would face a multiple of this cost with a general-purpose LLM.

Two factors significantly affected real-world costs in our analysis. The first is prompt caching. Our system prompt and output schema are the same for every note, and so caching significantly reduced costs. For Claude, caching has to be specifically enabled; when we enabled caching, Sonnet’s cost fell by 47%, from $30.70 to $16.32 for the 3,000 notes. The GPT-6 models cached prompt content automatically, with 58% to 73% of their input tokens served from cache. Haiku could not use prompt caching for our case, because it requires a prefix of at least 4,096 tokens and our prompt and schema come to about 2,650 tokens.[74] The second is the size of the request. Jev has the lowest price per input token in the study, $0.042 per million, and charges nothing for output, but each request carries about 34 questions with every answer option written out. That comes to roughly 15,000 input tokens per note, about eight times what the GPT-6 models sent and four to six times what the Claude models sent, in addition to the note (the note itself is only a small portion of the Jev input payload). As a result, Luna cost three to five times less than Jev despite Jev’s lower per-token pricing.

Latency, more than cost, is what decides whether a model can be used inside the EHR. IBM’s work on response time found that productivity rises sharply once a system answers in under a second, and that the gain from moving a response from three seconds into the sub-second range is large enough to change how people work.[81] Clinicians will not wait several seconds for an alert to appear. Jev answered in a median of 0.45 to 1.21 seconds. The other four models we tested took 1.7 to 3.5 seconds. To support use of these models, we would need to evaluate the notes in advance and cache the results. With Jev, the extraction could plausibly happen during the CDS evaluation itself, while the rule is being evaluated, which is a different and much simpler design. Speed also matters for cost, because it determines how a model can be used. A model that takes several seconds per note and costs about a cent per note has to be run in batch mode, ahead of time, on every note that might someday matter. Most of those notes will never be used by a CDS rule, so much of the money spent is wasted, and the extracted data are already out of date by the time the rule fires. Batch processing can also be difficult to use with identifiable data: at the time of our study, Anthropic’s discounted Message Batches API was not listed as eligible for use under its HIPAA agreement.[82] A model that answers in about a second and costs a fraction of a cent per note could instead be run just in time, for example, when a primary care visit is scheduled or opened for a patient aged 50 to 80 years whose structured smoking data are incomplete. For the roughly 100,000 VUMC primary care patients in that age range, reading 50 notes each would cost about $3,100 to $3,300 with Jev, compared with $19,000 to $34,000 with Sonnet. This would make LCS reminders, and many similar rules, more accurate and more current.

There are several possible explanations for Jev’s errors in the complex condition, and most of them point to fixable problems. First, the largest group of errors came from our own question design: Jev answered questions about each smoking period independently, and when a note described a patient who quit and restarted, it usually reported a spurious third period that our code added in. Second, some patterns, such as a quit date defined by a grandchild’s current age, were simply missing from the question set. A structured-judgment model can only choose among the options it is given, so its question set has to anticipate the ways information is documented. This is a form of prompt engineering, and it needs to be tested against real notes. Third, Jev’s answers include probabilities, which the vendor describes as calibrated, and a confidence value, and its “other or unclear” options gave it a way to decline. We did not use confidence here, but it is returned to the calling code, so a rule could treat a low-confidence answer as missing data rather than acting on it. Finally, Jev cannot generate text or explain its answers, which makes it unsuitable for some tasks but also removes a common failure mode of generative models: it cannot invent a number that does not appear among its options. As the notes about restarted smoking showed, however, code that combines its answers can still produce a wrong value.

The general-purpose LLMs had different weaknesses. Haiku, the less expensive Claude model, made arithmetic errors (particularly in converting an age at quitting to a calendar year and in adding up smoking periods) that led to the largest number of eligible patients classified as not eligible of any system (10, 5, and 13 in the three conditions). These are the most consequential errors for LCS, because a patient who is incorrectly classified as not eligible will not be prompted for screening. Sonnet made very few errors, but it cost 6 to 10 times as much as Jev. In our study, most of the additional accuracy that the higher price bought came from the hard cases. The two OpenAI models were at least as accurate as their Claude counterparts on every field on the complex notes. Luna showed the same weakness as Haiku, in arithmetic over several smoking periods, but less often, and Sol was the only system other than Sonnet that made fewer than five errors of any kind in the complex condition.

Our findings are consistent with prior work showing that LLMs can extract smoking history from notes with high accuracy.[27, 28] Luo et al. found that generative LLMs exceeded 96% accuracy across seven smoking variables and deployed one of them to more than 79,000 notes.[27] Dávila-García et al. found that a lightweight open-source LLM was non-inferior to cTAKES for smoking status, with inference times of 2.5 to 14.5 seconds per note.[28] In our prior work at VUMC, combining NLP-extracted smoking information with structured data identified 73.8% more patients eligible for LCS than structured data alone.[42] Our study adds two things to this literature: a direct comparison of accuracy with cost and speed, and a shareable benchmark on which other models can be compared.

It is worth noting that Jev, and to a lesser extent Haiku, made more errors on the template-generated notes than on the LLM-written notes, even though the LLM-written notes were longer and messier. The template generator expressed some facts in terse shorthand, such as “x39y”, while the LLM writer usually spelled the same facts out in full sentences, even when the rest of the note was messy. Real notes contain a great deal of shorthand, so this suggests that LLM-written synthetic notes may be easier than real notes, and that a benchmark should include both kinds of text.

To support broader adoption and further testing, several advances are needed. First, a model of this type needs to be available in a form that can be used with patient data: deployed locally, or hosted under a BAA with appropriate data handling, which TypeSafe’s documentation does not currently describe.[64] Second, the model and our question set need to become more accurate, particularly for complex histories, and they need to be validated on real notes. Third, it needs to be integrated into the EHR so that it can be called when a CDS rule is evaluated, with its answers and confidence available to the rule logic.

Our study has several strengths. The benchmark was designed for a specific, high-value CDS use case, and its three conditions separate the effect of messy text from the effect of complex arithmetic. Because the reference labels were generated before the notes, they reflect exactly what each note says, and because the notes are synthetic, the benchmark, code, and results can be shared openly for other models to be compared. We froze both systems before the harder conditions were created, adjudicated every disagreement, and measured cost and speed along with accuracy.

It also had some important limitations. First, the notes are synthetic. Although the LLM-written notes are realistic, they are not real, and as noted above they may be easier than real notes in important ways. The LLM-written notes were also written by a Claude model, which may favor the Claude systems, although the systems were frozen before the notes were created. Second, the first author designed both the generators and the TypeSafe question set, and the basic condition’s documentation patterns were known when the question set was written; the messy and complex conditions were designed after both systems were frozen to reduce this advantage. The blinded review and adjudication of the reference labels were also performed by an LLM rather than by clinicians. Third, we used one run of each system at one point in time, and, as the four days between our two sets of runs showed, that point moves. Model versions, prices, rate limits, and latencies change, and Jev’s latency varied between runs. The two OpenAI models were also released after the benchmark was published, so they had the advantage of being newer, although their knowledge cutoffs precede its publication. Fourth, our eligibility framework was simplified: it did not consider other factors in screening decisions, such as comorbidities, life expectancy, prior screening, or shared decision-making. Finally, we tested one structured-judgment model and four general-purpose LLMs, and other models in either class may perform differently.

## 5 Conclusion

A low-cost structured-judgment model extracted smoking histories from synthetic clinical notes with accuracy similar to general-purpose LLMs for routine documentation, and it was the only system we tested that answered in under a second, but it was less accurate for complex histories and, within four days of our first runs, it was no longer the cheapest option. Structured-judgment models still have advantages for this task: they answer faster, they cannot report a value that was not among their options, and they return a confidence that a rule can act on. General-purpose LLMs may close those gaps, particularly if their latency falls, and the cost gap has already closed. Models of this type, from either class, could make real-time, note-based CDS practical for LCS and many other uses. Before they are used in care, they should be validated on local notes and deployed in a HIPAA-compliant manner. They should also be re-evaluated often. Both the cheapest and the most accurate system in our study changed while we were writing it, and they will change again.

## Data Availability

All data produced are available online at https://github.com/vclic/smokingbenchmark and https://github.com/vclic/smokingeval.

https://github.com/vclic/smokingeval.

## Data and code availability

The benchmark (3,000 notes and reference labels) is available at https://github.com/vclic/ smokingbenchmark under a Creative Commons Attribution 4.0 International license. The code used to run and score each system, including the prompts, schemas, and TypeSafe question sets and the SHA-256 hashes recorded when they were frozen, is available at https://github.com/ vclic/smokingeval. The predictions and run metadata for all fifteen runs, and for the uncached Claude Sonnet 5 runs, are included in that repository.

## Use of generative AI

Claude (Anthropic) was used to help write the code for the benchmark generators and the evaluation, to generate the LLM-written notes as described in the Methods, to perform the blinded abstraction and adjudication of reference labels described in the Methods, and to help draft and edit this manuscript. The authors reviewed and take responsibility for the content of the manuscript.

## Author contributions

Adam Wright conceived and designed the study, developed the benchmark, conducted the analysis, and drafted the manuscript. Siru Liu and Aileen Wright contributed to the interpretation of the findings and critically revised the manuscript. All authors approved the final version. Adam Wright takes responsibility for the integrity of the data and the accuracy of the analysis.

